# The risk of transfusion transmissible infection in a civilian walking blood bank using rapid diagnostic tests: A modeling study

**DOI:** 10.1101/2025.10.21.25336343

**Authors:** Neil Thivalapill, Zhaohui Geng, Nikathan Kumar, Nobhojit Roy, Bopaya Bidanda, Nakul Raykar

## Abstract

**Background:** Civilian walking blood banks (WBBs) transfuse fresh whole blood from mobilized donors, screened using rapid diagnostic testing (RDT) for transfusion transmissible infections (TTIs), to preserve life when banked blood is unavailable. However, concerns regarding TTI risk using a RDT process instead of a more traditional, laboratory-based test persist. We aimed to understand the marginal risk of TTI using an RDT-based strategy compared to a laboratory-based test through development of a simulation model and accompanying online tool.

**Study Design and Methods:** We modeled expected TTIs per 100,000 donations from initial collection to transfusion and seroconversion. Parameters included TTI prevalence, donor risk-stratification, efficacy of stratification tools, TTI testing rates, platform test performance, and probability of seroconversion.

**Results:** A baseline TTI prevalence of 1% (95% CI: 0.25%, 1.75%) resulted in 56 TTIs (95% CI: 23, 91) when the RDT sensitivity was 90% (95% CI: 88%, 92%), 30 TTIs (95% CI: 12, 52) when the RDT sensitivity was 95% (95% CI: 93%, 97%), and 12 TTIs (95% CI: 4, 23) when the RDT sensitivity was 99% (95% CI: 97, 100%) per 100,000 donations. Compared to lab-based testing, 15,351 donations would need to be made under a high-sensitivity RDT testing strategy in order to incur one additional TTI.

**Discussion:** In a simulated WBB model, modern RDT platforms demonstrated favorable test characteristics, with low absolute rates of TTI, particularly when low-risk donors are selected. These findings support WBB implementation as an emergency transfusion strategy in settings lacking banked blood.

## Introduction

Approximately 40% of deaths worldwide are due to bleeding.^1^ Blood for transfusion, however, remains scarce and almost all low-and-middle-income countries experience blood deficits.^2,3^ Compounding this challenge is the lack of blood-banking infrastructure in most of the world, resulting in extensive blood deserts.^2,3^ Even high-income countries have blood deserts, especially in rural areas which face limited access to blood. Mass casualty events, large scale combat operations, natural disasters, and supply chain disruptions put even the best resourced blood systems at risk of being overwhelmed and turning into a temporary blood desert.

One strategy to address the lack of banked, screened blood available for transfusion in emergencies is a walking blood bank (WBB).^4^ The basic tenets of a WBB consist of rapid mobilization of donors, point-of-care testing for blood typing and transfusion transmissible infections (TTIs) (e.g., human immunodeficiency virus (HIV), viral hepatitis (HBV/HCV), and syphilis), and whole blood transfusion instead of fractionating blood into its components.^5–7^ Organizers of WBBs may choose to alter any, or all, of these steps to suit the urgency and demands of their circumstances.^1–3^

WBBs have been used for decades to save lives in battlefield settings and rural medical practices. However, concerns about the effectiveness of rapid diagnostic testing (RDT) for detecting TTIs have limited wider adoption. These concerns center on whether RDT is as reliable as laboratory-based methods like enzyme immunoassay (EIA) or nucleic acid amplification testing (NAAT), particularly in regions where TTIs are common.

It is important for clinicians and blood and health systems administrators to understand the risk of TTIs in their context to inform decisions on employing an RDT-based, WBB emergency process when banked blood is unavailable. Theoretical, predictive models can provide important real-world insights.^4^ We aimed to create a predictive model of TTI risk through the blood supply using an RDT-based approach and understand the marginal risk of TTI transmission compared to an EIA or NAAT-based approach. Additionally, we aimed to consolidate these risk calculations into a tool that could be used by clinicians and health system administrators to input parameters specific to their context and make risk stratification decisions regarding WBB operation.

## Study Design and Methods

We built a mathematical chance model to incorporate the myriad factors that can influence infection transmission through transfusion processes. The model can be accessed at https://tti-testing-strategy.herokuapp.com/.

### Model Construction

Numerous parameters impact TTI risk in the setting of a WBB. These include baseline prevalence of the infection in the general population, the rate of risk-stratifying donors through donor questionnaires, the efficacy of risk-stratification which ultimately determines the prevalence of the infection in the risk-stratified donor pool, the rate of screening donor products, the choice of testing strategies and their associated characteristics/performance, and ultimately the risk of seroconversion. Due to historical inconsistency of the term “donor screening,” we use “donor risk-stratification” to refer to the process of using a donor questionnaire to reduce the likelihood of contribution from high-risk donors, and “blood product screening” as the process of testing collected blood for TTIs using RDT, EIA, or NAAT.

To clarify the role that each of these choices and parameters play in a hypothetical WBB, we defined a process map that begins with donation of blood products and ends with transfusion in a recipient vein (Figure 1). Each node corresponds to a parameter or a stakeholder-defined choice and is explained further below. Test case values for all nodes are summarized in Table 1.

**Figure 1:**
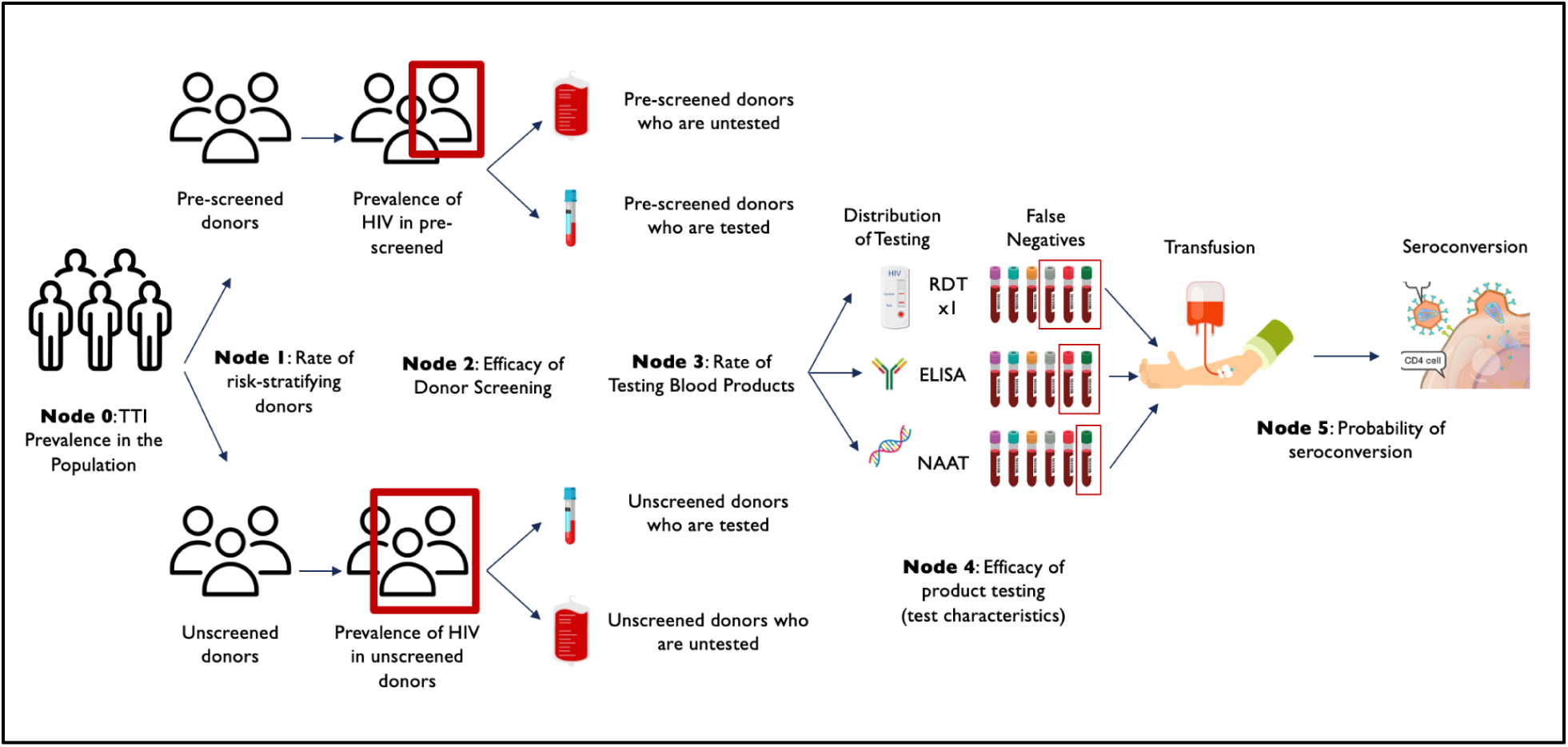
Process Map. The process map describes the flow of blood products beginning with donation and ending with transfusion into the recipient vein.

**Table 1.** Parameters in Our Test Case Simulation-Based TTI Modeling.

|  |  |  |
| --- | --- | --- |
| Node 0 | Low Population Prevalence | 1% (0.25%, 1.75%) |
|  | Medium Population Prevalence | 5% (4%, 6%) |
|  | High Population Prevalence | 10% (8%, 12%) |
| Node 1 | Donor Risk Stratification Screening Rate | 100% (100%, 100%) |
| Node 2 | Donor Risk Stratification Screening Efficacy | 50% |
| Node 3 | Product Screening Testing Rate Among Risk Stratified | 99% (98%, 100%) |
|  | Product Screening Testing Rate Among Not Risk Stratified | 99% (98%, 100%) |
| Node 4 | Product Screening Testing Efficacy - Sensitivity of RDT | Low - 90% (88%, 92%)<br>Mid - 95% (93%, 97%)<br>High - 99% (97%, 100%) |
|  | Product Screening Testing Efficacy - Sensitivity of EIA | 100% (98%, 100%) |
|  | Product Screening Testing Efficacy - Sensitivity of NAAT | 100% (99.9%, 100%) |
| Node 5 | Seroconversion Rate | 90% (85%, 95%) |

RDT = rapid diagnostic test; EIA = enzyme immunoassay; NAAT = nucleic acid amplification test

### Node 0 - TTI Prevalence in the Population

At the beginning of this process map, a population of WBB donors, derived from the general population, proposes to donate blood to the WBB. This population has an underlying prevalence of several TTIs such as HIV 1 and 2, Hepatitis B and C, syphilis, malaria, Human T-lymphotropic virus (HTLV) 1 and 2, Epstein–Barr virus (EBV), and Cytomegalovirus (CMV). For the purposes of model simplicity and ease of interpretation, the model focuses on the calculation of risk from one hypothetical TTI at three different levels of prevalence: low rate: 2% (95% CI: 1%, 3%), moderate rate: 6% (95% CI: 5%, 7%), and high rate: 10% (95% CI: 9%, 11%). When considering the total risk of multiple TTIs in a donor population, we propose calculating the total risk as the simple sum total of the individual TTI risks. That is to say, the total TTI risk ≤ TTIA + TTIB + TTIC. We recommend this approach despite the almost certain presence of concomitant infections in some donors in order to produce the most conservative estimate (overestimate) of TTI risk.

### Node 1 - Donor Risk Stratification Rate

Node 1 indicates the stakeholder’s choice of the rate at which to risk-stratify these donors using a standardized donor questionnaire to reduce the risk of TTI in the collected blood units.^8^ For the test case, we specify a donor risk stratification rate of 100% (lower bound (LB): 100%; upper bound (UB): 100%) to reflect the likely near complete donor risk stratification that would occur in a hypothetical civilian WBB optimized to minimize TTI risk. Again, based on implementation specifics and risk tolerance, communities may decide on less stringent approaches.

### Node 2 - Donor Risk Stratification Efficacy

Intertwined with the rate of donor risk stratification is the efficacy of stratification. We specified that donor stratification in this case yields a 50% reduction in TTI risk from the standard population, which is a conservative estimate based on studies of efficacy of validated donor risk stratification tools, though this varies in the literature and is often as high as 80-90% reduction in risk.^9^ That is to say, if a risk stratification tool is used, it is 50% effective at averting donations from individuals who will donate a TTI-positive blood product and the TTI prevalence among the risk stratified pool may be considered as one-half of the TTI prevalence among the non-stratified pool. In the online model, we provide a sliding scale to allow users to specify the x-fold reduction in TTI prevalence that is a result of donor risk stratification with a range from x=1, indicating no efficacy of donor risk stratification, to x=20, indicating an efficacy of 95%.

### Node 3 - Product Screening Rate

From Node 2, products originate from either the risk-stratified or non-risk stratified pool, and at Node 3, stakeholders are now asked to specify the rate at which they screen both groups of donated products for TTIs using a testing platform (RDT, EIA, or NAAT). For ease of interpretation, the test case assumes that products from both groups of donors are tested at the same rate of 99% (LB: 98%, UB: 100%). However, the model is flexible and allows for users to specify different testing rates from these two potential donor pools. Given the potential higher prevalence of TTIs in the non-stratified donor pool, it is possible that some users may choose to specify a higher rate of donor testing for blood products that originated from these non-stratified donors compared to those products that originated from risk stratified donors, depending on local context.

### Node 4 - Product Screening Efficacy

Arriving at Node 4 in the process map, the donor has either been risk stratified or not using a donor questionnaire and the donated product has either been screened or not screened for TTIs. Node 4 represents the testing regimen specified for implementation by the stakeholder and the efficacy of that strategy. In the model, we offer three testing strategies: RDT, EIA, and NAAT. The model requests that the user specifies the sensitivity of each of these screening strategies given that the false negative rate, which is given as *1-sensitivity*, defines the source of TTI risk. The online model also provides a table of the estimated TTI risk under each screening strategy.

The testing characteristics of each platform (RDT, EIA, and NAAT) are known to vary with the type of infection, the geographical context, the presence of comorbid infections, and the acuity of infection. RDT platforms offer a timely, resource-conscious method of screening blood products but their widespread use in blood banking systems has been limited due to concerns of poor testing characteristics in earlier iterations. EIA platforms have historically addressed the testing characteristic/performance concerns of RDT but require significant centralized laboratory infrastructure to run these diagnostics and are simply unavailable in the world’s lowest resource settings. Similarly, while NAAT also demonstrates high sensitivity during early acute infections with a smaller “window period” of infection – the gap between when an individual is infected and laboratory-detectable levels of antibodies or antigen are present in their blood – NAAT implementation is limited in resource-constrained settings due to cost.^5^ The screening characteristics of RDT are set to three different levels in this case study to highlight the comparison: low RDT sensitivity: 90% (88%, 92%), moderate sensitivity: 95% (93%, 97%), and high sensitivity: 99% (97%, 100%). Also for the purposes of the case study, we specified high sensitivities for EIA (100%, LB: 98% UB: 100%) and NAAT (100%, LB: 99.9% UB: 100%) in order to provide the most conservative estimate of the number needed to harm (NNH) for TTI risk under an RDT-based strategy.

### Node 5 - Rate of Seroconversion

The final node in the process map represents the probability of seroconversion once an infected blood product is transfused into a recipient. The probability of seroconversion depends on several factors including the specific TTI, the pathogen load in the blood sample, and donor characteristics, such as CD4 receptor compatibility with HIV. Previous estimates suggest that the risk of seroconversion may range from 50% to 95% depending on these factors and, for the purposes of overestimating the risk of infection to produce a conservative estimate, we specify a seroconversion rate of 90% for the test case (LB: 85%, 95%).

### Model Simulation

We leveraged a discrete-event simulation technique to model the WBB system, as described above. At the beginning of the simulation, 100,000 donors are independently generated to enter the system and go through all the previously described nodes (decision points) based on their corresponding decision criteria and rates. The probability of each path at a given node can be specified by the user and values used for these nodes for our test case are listed in Table 1. The remaining variables are modeled as triangular random variables, whose distribution is specified by three variables: a mode and a lower and upper bound. Given the model is designed to produce values for a specific TTI, and this is influenced by both the test characteristics of RDT, EIA, and NAAT specific to that TTI, as well as the TTI prevalence in the population, the values used in the test case are specified in Table 2. TTI prevalence in the population was selected through a review of values reported for rural, low-resource regions in the world. As with the rest of the model, these values can be adapted to local context.

**Table 2.** TTI Prevalence, Respective Testing Characteristics, and TTI risk per 100,000 transfusions.

| **TTI Prevalence** | **RDT Sensitivity:**<br>**90% (88%, 92%)**<br>**Rate per 100,000**<br>**transfusions** | **RDT Sensitivity:**<br>**95% (93%, 97%)**<br>**Rate per 100,000**<br>**transfusions** | **RDT Sensitivity:**<br>**99% (97%, 100%)**<br>**Rate per 100,000**<br>**transfusions** |
| --- | --- | --- | --- |
| 1% (0.25%, 1.75%) | 56 (23, 91) | 30 (12, 52) | 12 (4, 23) |
| 5% (4%, 6%) | 281 (221, 347) | 153 (106, 204) | 59 (27, 97) |
| 10% (8%, 12%) | 561 (443, 693) | 306 (212, 409) | 118 (53, 194) |

RDT = rapid diagnostic test; TTI = transfusion transmitted infection

## Results

Final estimates consist of the number of TTIs resulting from the 100,000 independent and identically distributed copies of the donors moving through the system along with 95% confidence intervals. The parameters used in the test case are described in Table 1. Using these parameters, the sensitivities of the three testing strategies (RDT, EIA, or NAAT), prevalence, and NNH of an RDT-based strategy against EIA or NAAT-based strategies are reported in Table 2 and Table 3.

**Table 3.**
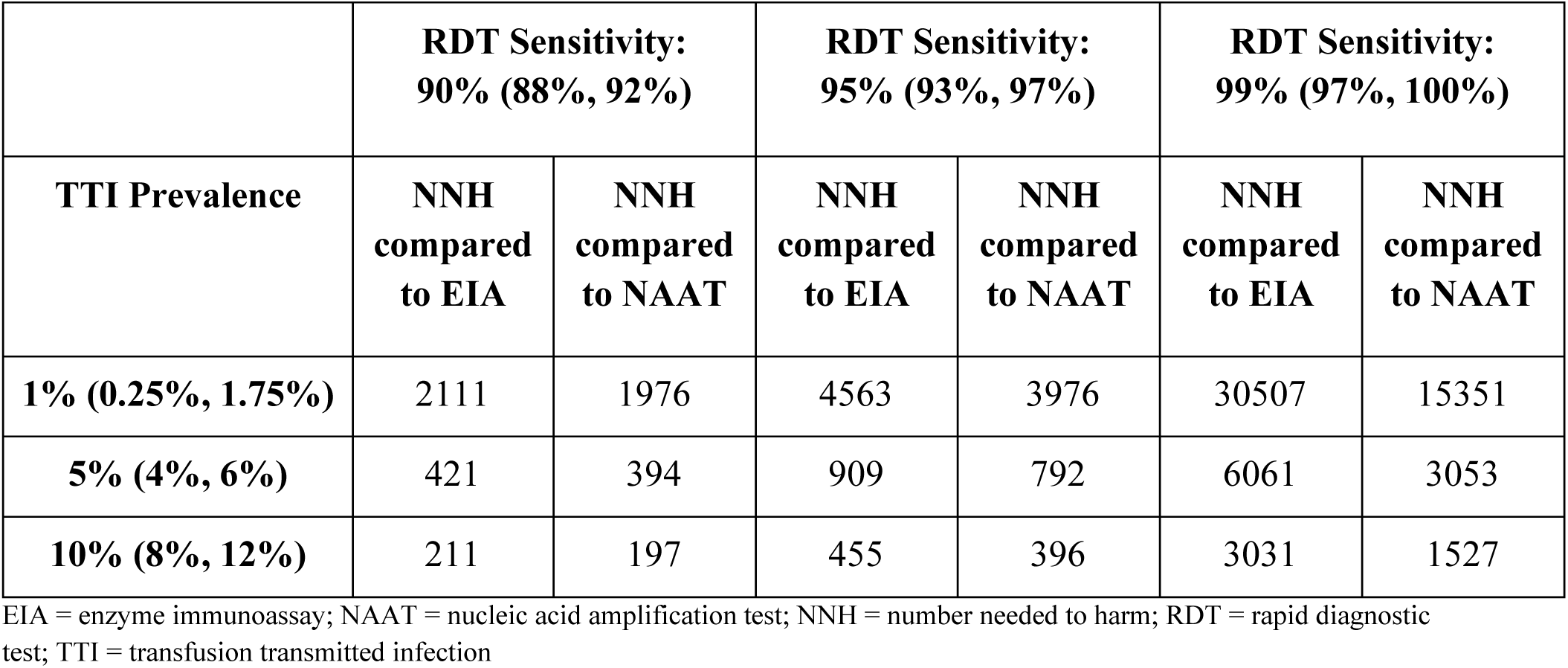
Number of transfusions needed prior to additional case of TTI transmission comparing RDT at various sensitivities to either EIA or NAAT.

For a theoretical TTI prevalence of 1% (0.25%, 1.75%) in the general population and an RDT sensitivity of 90% (95% CI: 88%, 92%), the associated rate of TTI was 56 per 100,000 transfusions (95% CI: 23, 91). This risk increased to 561 (95% CI: 443, 693) transmission events per 100,000 transfusions when the prevalence of the TTI increased 10-fold to 10% (95% CI: 8%, 12%) and decreased to 12 (95% CI: 4, 23) transmission events when RDT sensitivity was 99% (95% CI: 97%, 100%). As hypothesized, as TTI prevalence increased, the risk of transmission events increased and as RDT sensitivity increased, the risk of transmission events decreased.

Table 3 describes the number of transfusions needed before an additional transmission event occurred under an RDT vs. EIA strategy and under an RDT vs. NAAT strategy. Assuming a TTI prevalence of 1% and an RDT sensitivity of 90% (95% CI: 88%, 92%), the NNH to incur one additional transmission event under an RDT strategy compared to an ELISA strategy was 2,111 vs. 1,976 when compared to an NAAT strategy. The NNH when comparing RDT to EIA decreased to 211 when prevalence was increased to 10% (95% CI: 8%, 12%) and increased to 30,507 when RDT sensitivity increased to 99% (95% CI: 97%, 100%). For all TTI prevalence and RDT sensitivity scenarios, the NNH comparing RDT to EIA was higher than the NNH comparing RDT to NAAT.

## Discussion

In emergency situations where blood is needed and standard laboratory-screened blood is unavailable, transfusion of blood using RDT-screened blood carries a low risk of TTI transmission per donation. In a theoretical scenario with 1% TTI prevalence with high-sensitivity RDT (99%), an additional transmission would occur only once every 15,351 transfusions. In a worst-case scenario, with 10% prevalence of the TTI and using EIA as the standard, an additional transmission would occur once every 3,031 transfusions. These are conservative estimates that deliberately overestimate infection risk.

These data support growing calls in high- and low-resource contexts alike to establish backup RDT-based processes to prepare for emergency or disaster scenarios where blood demand outstrips available supply of EIA- or NAAT-tested blood, or where blood banks or blood testing infrastructure are incapacitated.^6–8^ The model provides clinicians, patient advocates, and policymakers with an opportunity to quantify the TTI risk and weigh it against the risk of death from blood unavailability. Health system authorities should carefully examine these results, adapt for their context, and determine the extent to which an RDT-based WBB strategy could save lives at an almost infinitesimal TTI transmission risk.

Three notable factors have contributed to challenging long-held dogma of RDT inferiority and flipped the paradigm in favor of RDT-based WBBs when banked blood is not available. First, the latest generation of RDT has improved dramatically in its reliability, sensitivity, and specificity compared to older versions, with test performance comparing well to EIA and NAAT testing. Manufacturer-reported sensitivities for RDTs are in the range of 98-99% (Supplemental Table 1). Second, the WBB approach has been operationalized by the US military with exceptional success, having transfused over 10,000 units of fresh whole blood in combat settings with excellent safety profiles. Even in the civilian sector, WBBs have been established to meet community blood needs where banked blood transport is unavailable.^10^ Third, there is growing recognition of the need for backup approaches for blood transfusion in the context of disaster or mass casualty events or supply chain disruptions where standard blood banking operations may cease, even in traditionally well-resourced contexts.

We provide a sensitivity analysis, in Figures 2 and 3, of how results may change with increasing population prevalence or with poorer RDT performance. In general, the risks of TTI seroconversion are higher in higher prevalence populations, as well as in RDT platforms with poorer sensitivities, which leads to a higher TTI seroconversion risk at any given prevalence. Population risks and prevalence of TTIs, however, expose all testing platforms to the same challenges, and these trends are consistent across testing platforms (including EIA and NAAT).

**Figure 2.**
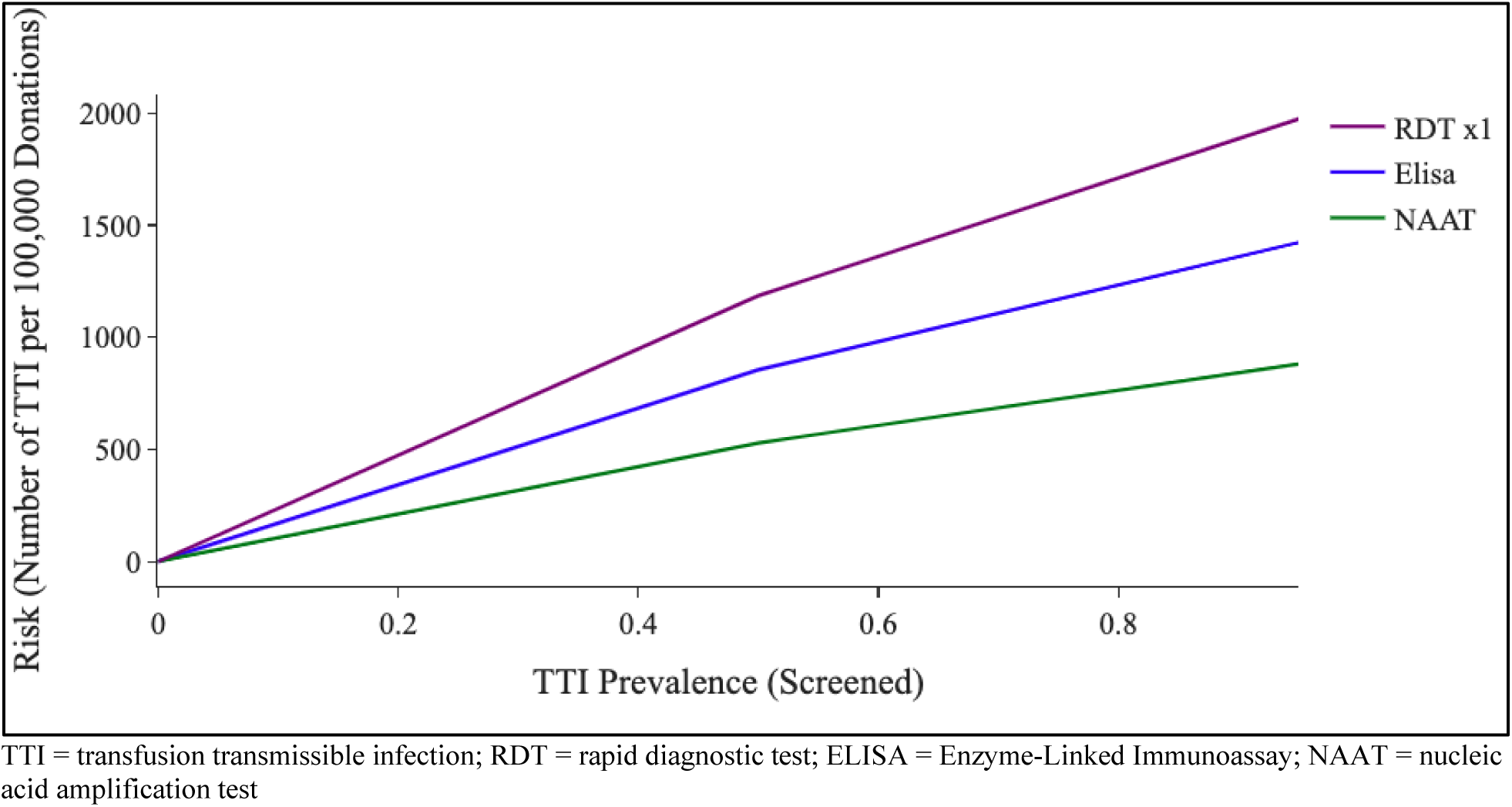
TTI Prevalence (Among Screened) vs. TTI Risk Under a High RDT Sensitivity Strategy.

**Figure 3.**
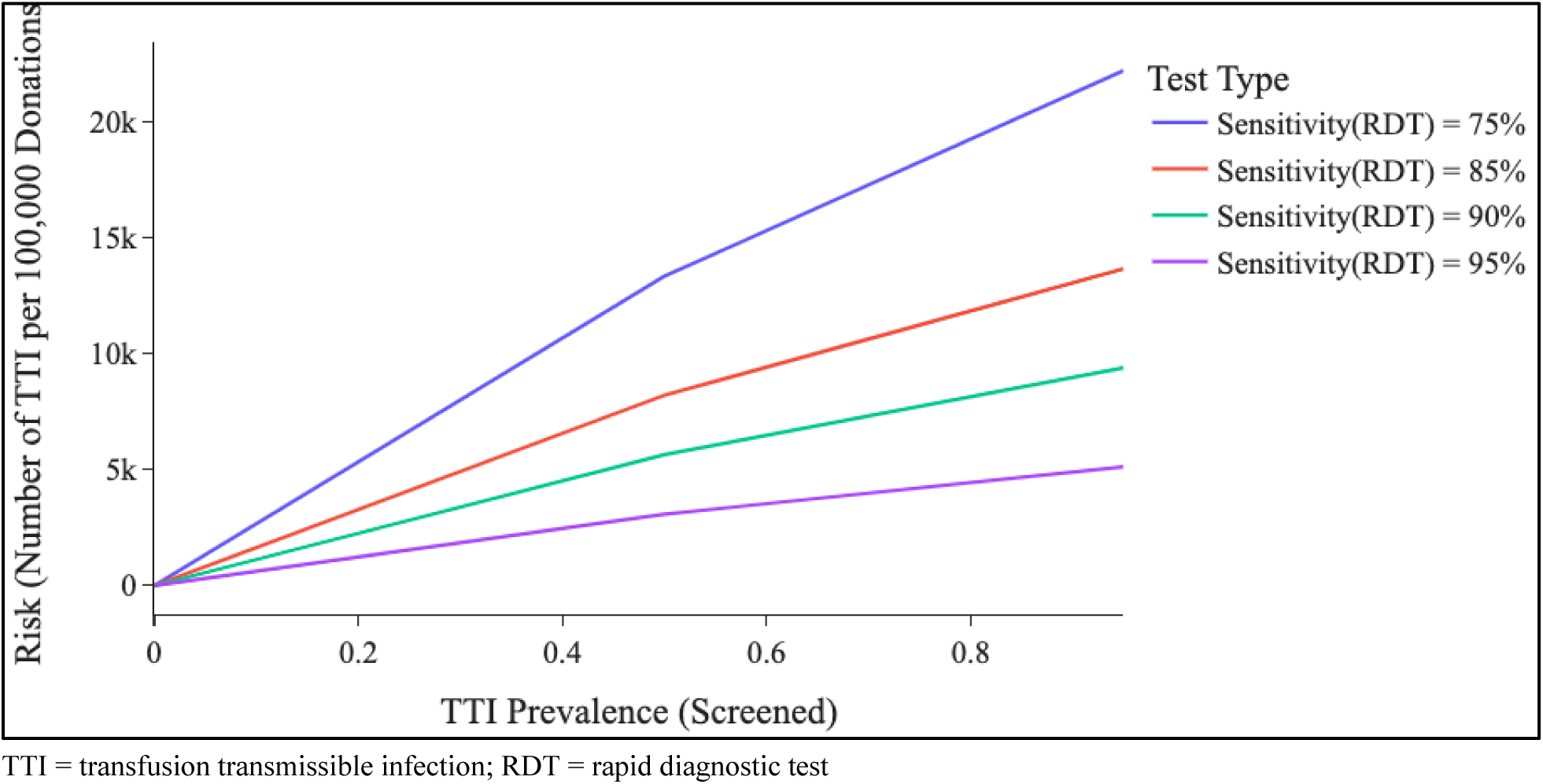
TTI Risk Under Various RDT Sensitivity Strategies.

It is important to note that the sensitivity of a test notes its ability to detect TTI when it crosses a threshold of detection, and that the greatest threat to false negatives in light of high test performance is the well-known ‘window period’ in the beginning of an infection where the viral or bacterial load is too low for detection.^9^ Testing for infection in the window period can lead to false negative results with any testing platform. Nonetheless, NAAT is known to have the shortest window period, superior to that of EIA and RDT.

There are important limitations to the model. This analysis makes specific assumptions that must be tailored and evaluated in the local context. First, at Node 1, the proportion of risk-stratified donors was set to 100%. Prior iterations of WBBs and emergency donor panels, particularly in the military, have relied on a group of pre-screened donors and specified that blood products of unscreened donors would be used only if pre-screened donors are unavailable.^4,11^ In dire circumstances, however, stakeholders may choose to screen fewer individuals in order to enlarge the donor pool. The model allows for flexibility to specify a lower donor screening rate. At Node 2, there is heterogeneity in the literature on screening efficacy, and multiple studies suggest that screening can result in a reduction of 95% or higher.^12^ We chose to use a *lower* efficacy of 50%, motivated by an effort to model a conservative scenario and again, we encourage stakeholders to leverage the flexibility of the model and use a donor screening efficacy that is appropriate for the local context. Similarly, we specified that nearly all (99%) blood products from both risk-stratified and non-risk-stratified donors would be tested for TTIs prior to transfusion. There are circumstances, for example, in military settings where donors are already pre-screened, where RDT was not used prior to transfusion. Given this manuscript was focused on an optimized RDT-based process for the civilian context, we chose to focus on the scenario where nearly all were tested. Finally, Node 5 represents the risk of seroconversion once an infected blood product is transfused. Given the extremely low rates of TTI events in modern blood banking, and the limited data available on seroconversion rates in humans, we assumed a 90% seroconversion rate for all TTIs. Previous studies have suggested that seroconversion may be as low as 40%, while other papers use a 75% seroconversion rate.^13–15^ Again, the choice to use a 90% seroconversion rate was motivated by the goal of modeling the upper bound of TTI risk but stakeholders using the calculator can specify a lower seroconversion rate.

This model assumes that RDT, EIA, and NAAT testing platforms all perform according to their package inserts and that the latest generation of each of these platforms is used. This renders EIA and NAAT performance as near exceptional. There may be heterogeneity in performance of all testing platforms based on the type of blood used for testing (capillary, whole blood, plasma), the conditions under which the tests are used, the prevalent comorbidities and co-infections, the reference standard used, as well as the operating conditions of the tests. There is great variability in the age of EIA and NAAT systems in current use worldwide. Indeed, the major benefit of this tool is that it can be adjusted to fit a large variety of environments. Finally, on a minor note, this is a model of independent donations and assumes that each independent donation leads to a single transfusion. This was chosen for simplicity of understanding the risk profile of obtaining blood from a single donor in an emergency transfusion scenario.

While this study compares the theoretical risks of RDT against EIA and NAAT, in reality, this is a false comparison in the decision-making lens of whether to mobilize a WBB. The risk of TTI transmission through an RDT-based process should be weighed against the *risk of death* from lack of transfusion as banked blood screened by EIA or NAAT is *unavailable*. Nonetheless, the comparison of RDT against EIA and NAAT is an important one that allows stakeholders to make informed decisions when EIA and NAAT blood is unavailable.

Using the latest generation RDT platforms and model parameters selected to approximate real-world scenarios, the risk of TTI can range from 12-561 TTIs per 100,000 donation-transfusions. Given the small-volume nature of WBBs, TTI risk in this setting is minimal and therefore should not be considered a limitation to WBB implementation in blood scarce settings. Further, the increased sensitivity of new RDT platforms results in an increased NNH when considering the risk of an additional TTI under an RDT-based strategy compared to an EIA-based strategy. We encourage stakeholders to leverage the online decision-support tool to estimate the TTI risk under their specific parameters.

We emphasize, however, that comparing RDT performance against an alternative of best-case scenario (EIA or NAAT) is ultimately a false comparison as risks for TTI transmission through an emergency WBB using RDTs should be compared against the alternative available in that situation – no transfusion. The risk of TTI transmission through an RDT process, then, should be weighed against the risk of death from lack of transfusion as traditionally banked blood is *unavailable*.

## Data Availability

All data produced in the present study are available upon reasonable request to the authors.

