## Supplemental Table 1 for "The risk of transfusion transmissible infection in a civilian walking blood bank using rapid diagnostic tests: A modeling study"

| Infection | Rapid Diagnostic Testing | Enzyme-Linked Immunoassay (EIA/ELISA) |
|---|---|---|
| HIV | Abbott Determine HIV-1/2 Ag/Ab: 99.9% (99.4%, 100.0%)[1] | Genscreen HIV Combo Ag/Ab EIA: 100%, (99.70%, 100%)[2] |
| Hepatitis B | Abbott ABON HbSAg Rapid Test: 99.13%[3] | Elecsys HBsAg II ECLIA: 100% (99.34%, 100%)[4] |
| Hepatitis C | Abbott Abon HCV Rapid Test: 99.53% (98.62%, 99.90%)*[5] | ORTHO HCV V 3.0 ELISA Test System: 100.0% (92.9%, 100.0%)[6] |
| Syphilis | Abbott Bioline Syphilis 3.0 Rapid Test: 99.3%[7] | Abbott ARCHITECT Syphilis CMIA: 100.0% (97.22%, 100%)*[8] |
